# APOL1 Monoallelic and Biallelic genotypes and CKD, Proteinuria and FSGS in African Americans from the Vanderbilt Biobank (BioVU)

**DOI:** 10.1101/2025.11.05.25339478

**Authors:** Steven W. Clapp, Jefferson L. Triozzi, Zhihong Yu, Fatih Mamak, Soumya Vavilala, Lee Wheless, Robert Corty, Hua Chang Chen, Otis D. Wilson, Guanchao Wang, Alexander Bick, Lide Han, Douglas Ruderfer, Cassiane Robinson-Cohen, Ran Tao, Adriana M. Hung

## Abstract

**Background:** African Americans are at increased risk for chronic kidney disease (CKD) in part due to Apolipoprotein L1 gene (APOL1) high-risk genotypes. Recently, a study from West Africa reported an association between monoallelic genotypes and risk of CKD and focal segmental glomerulosclerosis (FSGS). We here study the association of biallelic and monoallelic genotypes with CKD, proteinuria, and FSGS in participants in a hospital-based cohort in the Southeastern United States.

**Methods:** We conducted a case-control study of African Ancestry participants from the Vanderbilt University Medical Center Biobank (n=23, 857). The primary outcome was CKD defined as: persistent GFR < 60 ml/min, end stage kidney disease (ESKD), biopsy-proven FSGS or urine protein-to-creatinine ratio of >700 mg/g or albumin-to-creatinine ratio of >420 mg/g. Secondary outcomes were proteinuria and FSGS outcomes separately. The primary exposure was *APOL1* monoallelic genotype (1 copy of a risk allele versus none). APOL1 biallelic genotypes was studied as a secondary exposure. Sequential logistic regression models were performed adjusting for potential confounders.

**Results:** Among 23,857 participants, 5,784 had CKD, 1,533 had proteinuria, and 80 had biopsy-proven FSGS, 44.5% had one risk allele (monoallelic) and 13.6% had two risk alleles (biallelic). Biallelic carriers had higher odds of CKD than those with one or no risk alleles (adjusted odds ratio (*aOR*), 1.72; 95% confidence interval [CI], 1.57-1.89), proteinuria *aOR* = 2.02 (95% CI, 1.77-2.31), and FSGS *aOR* = 17.48 (95%CI 10.53-29.02). Monoallelic carriers (G0/G1 or G0/G2) had a small increase in odds of CKD (*aOR =* 1.08, 95% CI 1.00-1.16, p=0.04), which was driven by the G0/G2 genotype (*aOR =* 1.11, 95% CI 1.01-1.23, p=0.03). Monoallelic carriers (G0/G1 or G0/G2) had higher odds of proteinuria (*aOR* = 1.23; 95% CI, 1.09-1.39) and G0/G1 had higher odds of FSGS (14 cases) (*aOR* = 2.83; 95% CI, 1.18-6.79).

**Conclusions:** In our study, monoallelic APOL1 genotypes were associated with 23% higher odds of proteinuria and 3-fold higher odds of FSGS for G0/G1. A modest increase odd for CKD of 8%, which may reflect CKD phenotypic heterogeneity. Our study supports the observations from a West Africa cohort in a US based cohort and adds the association of APOL1 monoallelic genotypes with proteinuria.

## Introduction

In the United States, African Americans have a four-fold increase in the risk of ESKD. Although some of these differences can be attributed to socioeconomic factors and other comorbidities, previous studies have identified genetic variants in apolipoprotein L1 (*APOL1*) that partially explain this excess risk^1–3^.

The two *APOL1* high-risk (HR) variants are termed G1 and G2 and are common in individuals of West African descent as they confer resistance to lethal *Trypanosoma brucei Rhodesiense* infections, which is a pathogen that was endemic to West Africa^4, 5^. APOL1 is a circulating component of the innate immune system that protects against parasitic infection by forming nonselective cation channels in the plasma membrane leading to colloid osmotic swelling and eventual parasite lysis. Although the genetic link between *APOL1* risk variants and kidney disease is well-established, the mechanism by which *APOL1* risk variants induces kidney injury remains uncertain with multiple mechanisms considered^6^. However, the scientific community concur that APOL1 mediated kidney disease is primarily a podocytopathy with proteinuria being a key clinical manifestation that may precede the development of FSGS or irreversible kidney damage^7, 8^

Until recently it was considered that two copies of high-risk variants were required to acquire the excess risk for kidney disease, homozygous for G1, homozygous for G2 or compound heterozygous G1/G2^1^. However, a recent study from West Africa challenged this scientific premise by demonstrating that one copy of G1 or G2 (monoallelic carrier) was sufficient to confer risk of kidney disease or focal segmental glomerulosclerosis (FSGS), albeit at a lower risk^8^. These findings have tremendous implications for precision nephrology and clinical translation, given the high allele frequencies for the high risk variants globally^9^. The associations of monoallelic carriers with CKD need to be replicated in the United States and further understood regarding population differences and environmental factors that could have determined the observed differences.

In our current study, we evaluated whether there is an association between monoallelic carriers and CKD compared to the known associations for biallelic carriers. We also study the association of monoallelic carriers and our secondary outcomes, which include proteinuria and FSGS.

## Methods

### Study Design and population

We conducted a case-control study of individuals of African ancestry participating in the Vanderbilt University Medical Center’s, EHR-linked biobank (BioVU)^14^. We included those that had whole genome sequencing data from the Alliance for Genomic Discovery (AGD)/BioVU (n=27,782)^10, 11^. The source of phenotypic data for this study was the de-identified mirror of the EHR, known as the BioVU Synthetic Derivative. Patients were included if they were at least 1 year old and had at least one creatinine value if normal value. In individuals who had an abnormal creatinine value, a second value with a corresponding eGFR < 60 was required 90 days apart to meet criteria for CKD. Others could meet CKD criteria with documented proteinuria, biopsy-proven FSGS, or other CKD criteria were required (see below). We excluded individuals with other forms of monogenic kidney disease such as Alport (n = 5) and ADPKD (n = 94), individuals with biopsy-proven diabetic kidney disease (n=5), decompensated cirrhosis (n = 394), end stage heart failure with or without transplant (n = 104), lung transplant (n = 45), and liver transplant (n = 119) or sickle cell disease (n=240). These criteria were selected based on a recent publication evaluating the same association^8^. Flow chart for the creation of the analytical cohort are presented in **Figure 1**. The VUMC Institutional Review Board oversees BioVU and approved this project.

**Figure 1.**
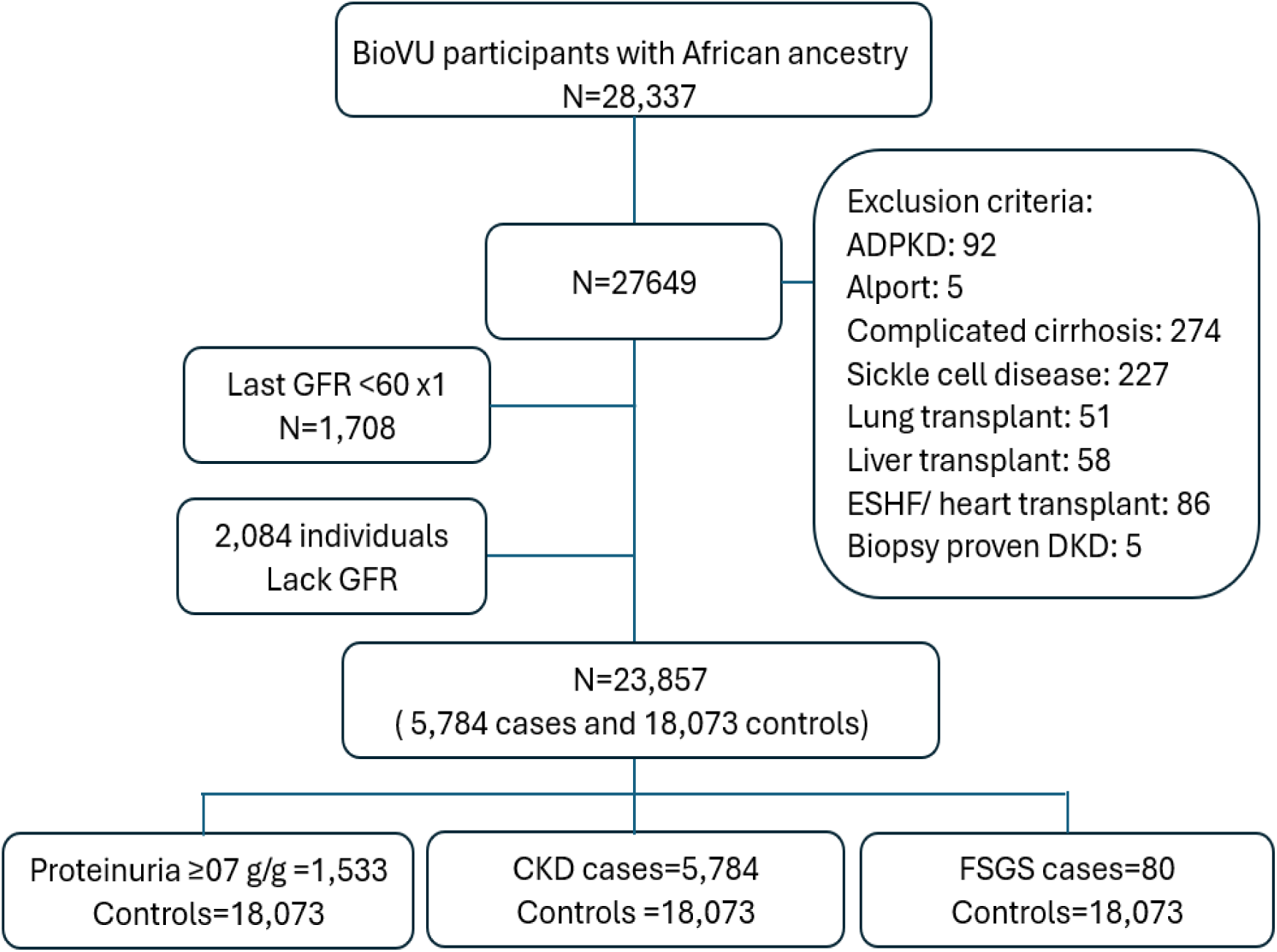
Flow chart for the study cohort

### Main exposure

The primary exposure was *APOL1* monoallelic genotype (1 copy of a risk allele versus none). APOL1 biallelic genotypes was studied as a secondary exposure. *APOL1* high-risk variants G1 (rs73885319 p.S342G; rs60910145 p.I384M) and G2 (rs71785313, a 6–base pair deletion that removes amino acids N388 and Y389) were directly genotyped on DNA that was extracted from whole blood. Participants were defined as high risk or biallelic if they had 2 high risk alleles: homozygotes for G1/G1, homozygotes for G2/G2, or compound heterozygotes for G1/G2. All the other participants were characterized as low risk. Monoallelic carriers included individuals that were G1/G0 or G2/G0. Each monogenic genotype was also studied following the analysis described by Gbadegesin et al.^8^ All genotypes were in Hardy-Weinberg equilibrium and present at frequencies comparable to prior reports and in population databases.

### Outcomes

#### Primary outcome

The primary outcome was CKD, defined as 1) estimated glomerular filtration rate (eGFR) <60 ml/min/1.73m^2^ on two occasions apart at least by 90 days, 2) two ICD-9 or ICD-10 codes (9th and 10th revisions of International Classification of Diseases) in at least two outpatient visits or one inpatient hospitalization (**supplemental table 1**), 3)) biopsy-proven FSGS or 4) proteinuria by protein to creatinine ratio (PCR) > 700 mg/g, albumin to creatinine ratio > 420 mg/g, code for nephrotic syndrome or code for persistent proteinuria. People with no kidney disease, documented GFR> 60 ml/min and no known proteinuria, were included as controls.

#### Secondary outcomes

Secondary outcomes included individual outcomes of 1) proteinuria defined as a PCR > 700 mg/g or an albumin to creatinine ratio > 420 mg/g. or 2) biopsy-proven FSGS, as determined by the biopsy report. The outcome cut-offs were chosen based on the threshold identified by the PARASOL initiative and replicated RaDar^12^ where protein: creatinine ratio >700 mg/g represented a cutoff associated with higher hazard of kidney failure.

### Sensitivity and exploratory analyses

<u>Sensitivity analyses</u> included 1) the highest PCR recorded as a continuous trait in individuals in whom measurements were available without a cutoff and 2) an additive model of inheritance, which assumes that each allele in the genotype adds effect on the phenotype (no risk, intermediate risk, full risk) for the primary and secondary outcomes.

#### Covariates

Characteristics reported include demographics, comorbidities, characteristics, laboratory tests, and medications (**Supplemental Table 1**). Ancestry determination was genetically inferred using global ancestry estimation. Admixture proportions were inferred by the Alliance of Genetic Discovery using SCOPE, which estimates an individual allele frequency matrix through latent subspace estimation and then decomposes the estimated matrix into ancestral allele frequencies and admixture proportions. Individuals were assigned to the ancestry cluster for which the proportion exceeded 0.5 (majority of ancestry proportion).

#### Statistical analysis

The primary exposure variable was *APOL1* monoallelic genotypes one risk variant (G0/G1 and G0/G2) compared to none (G0/G0). We also studied high-risk group defined by the presence of 2 risk variants versus 1 or none (recessive model of inheritance-biallelic group) for face validity, and explored each individual genotype. Sequential logistic regressions were used to study the association of the exposure and the outcomes of interest. The primary analysis was the minimally adjusted model, which adjusted for age, sex, diabetes status (regardless of its documented timing to kidney disease) and 3 principal components of ancestry. The decision to adjust for diabetes rather than stratification was because in many, diabetes developed after CKD onset. The fully adjusted model, adjusted for BMI, mean arterial pressure closest to the GFR value or the diagnosis of CKD, number of antihypertensives, HIV status, renin angiotensin aldoterone system inhibitors (RAASi), and SGLT2 inhibitors. Proteinuria was not treated as a covariate because it is included in the definition of CKD and it is one of the secondary outcomes. The unadjusted model is presented for informational purposes. For FSGS, given the small number of cases, the association was explored using the Firth logistic regression method to reduce the bias in maximum likelihood estimates of coefficients.

## Results

### Study Cohort and participants characteristics

We studied 23,857 BioVU participants of African ancestry, directly genotyped for APOL1 risk variants, and that statisfied our entry criteria (**Figure 1**). Baseline cohort characteristics by APOL1 genotype risk-group were summarized (**Table 1**). 61.8% (n=14,736) were Females. The mean age was 45 years (SD 19.6). In total, 5,784 (24%) had CKD, from whom 906 received a kidney transplant. Seventy-two patients had biopsy-proven FSGS. 6,111 (25.6%) had diabetes, and from those with diabetes, fifty percent had CKD (n=3044).

**Table 1.**
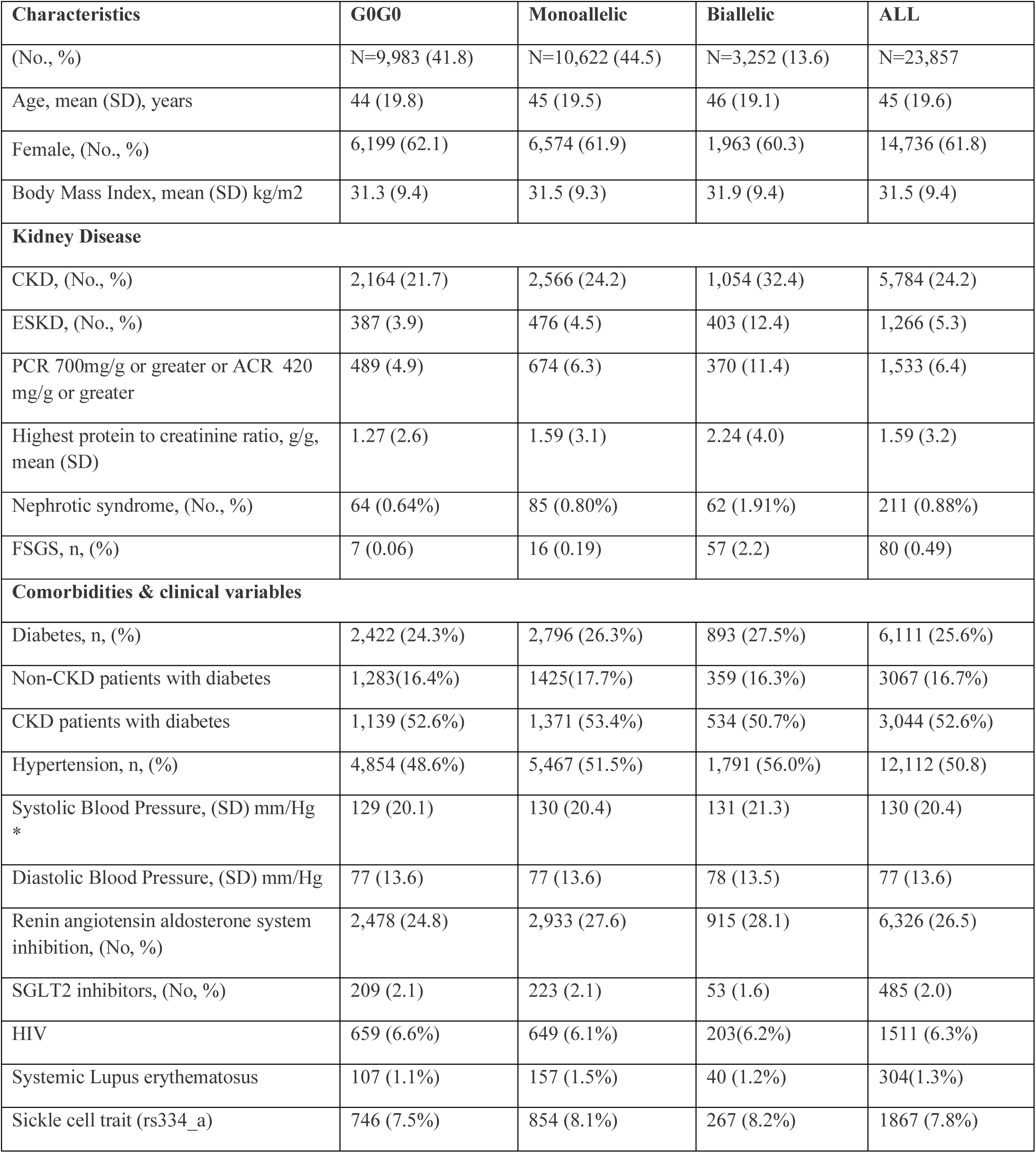
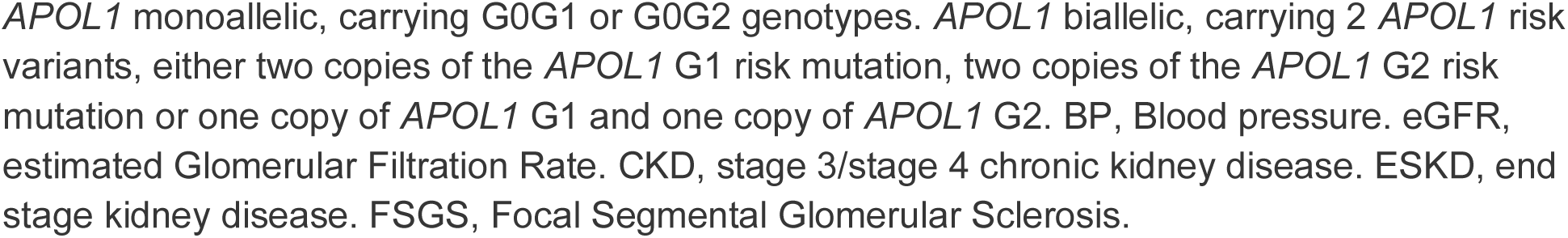
Clinical characteristics among 23,836 BioVU participants in the APOL1 Mediated Kidney Disease Cohort.

| Characteristics | G0G0 | Monoallelic | Biallelic | ALL |
| --- | --- | --- | --- | --- |
| (No., %) | N=9,983 (41.8) | N=10,622 (44.5) | N=3,252 (13.6) | N=23,857 |
| Age, mean (SD), years | 44 (19.8) | 45 (19.5) | 46 (19.1) | 45 (19.6) |
| Female, (No., %) | 6,199 (62.1) | 6,574 (61.9) | 1,963 (60.3) | 14,736 (61.8) |
| Body Mass Index, mean (SD) kg/m<sup>2</sup> | 31.3 (9.4) | 31.5 (9.3) | 31.9 (9.4) | 31.5 (9.4) |
| **Kidney Disease** |  |  |  |  |
| CKD, (No., %) | 2,164 (21.7) | 2,566 (24.2) | 1,054 (32.4) | 5,784 (24.2) |
| ESKD, (No., %) | 387 (3.9) | 476 (4.5) | 403 (12.4) | 1,266 (5.3) |
| PCR 700mg/g or greater or ACR 420 mg/g or greater | 489 (4.9) | 674 (6.3) | 370 (11.4) | 1,533 (6.4) |
| Highest protein to creatinine ratio, g/g, mean (SD) | 1.27 (2.6) | 1.59 (3.1) | 2.24 (4.0) | 1.59 (3.2) |
| Nephrotic syndrome, (No., %) | 64 (0.64%) | 85 (0.80%) | 62 (1.91%) | 211 (0.88%) |
| FSGS, n, (%) | 7 (0.06) | 16 (0.19) | 57 (2.2) | 80 (0.49) |
| **Comorbidities & clinical variables** |  |  |  |  |
| Diabetes, n, (%) | 2,422 (24.3%) | 2,796 (26.3%) | 893 (27.5%) | 6,111 (25.6%) |
| Non-CKD patients with diabetes | 1,283(16.4%) | 1425(17.7%) | 359 (16.3%) | 3067 (16.7%) |
| CKD patients with diabetes | 1,139 (52.6%) | 1,371 (53.4%) | 534 (50.7%) | 3,044 (52.6%) |
| Hypertension, n, (%) | 4,854 (48.6%) | 5,467 (51.5%) | 1,791 (56.0%) | 12,112 (50.8) |
| Systolic Blood Pressure, (SD) mm/Hg * | 129 (20.1) | 130 (20.4) | 131 (21.3) | 130 (20.4) |
| Diastolic Blood Pressure, (SD) mm/Hg | 77 (13.6) | 77 (13.6) | 78 (13.5) | 77 (13.6) |
| Renin angiotensin aldosterone system inhibition, (No, %) | 2,478 (24.8) | 2,933 (27.6) | 915 (28.1) | 6,326 (26.5) |
| SGLT2 inhibitors, (No, %) | 209 (2.1) | 223 (2.1) | 53 (1.6) | 485 (2.0) |
| HIV | 659 (6.6%) | 649 (6.1%) | 203(6.2%) | 1511 (6.3%) |
| Systemic Lupus erythematosus | 107 (1.1%) | 157 (1.5%) | 40 (1.2%) | 304(1.3%) |
| Sickle cell trait (rs334_a) | 746 (7.5%) | 854 (8.1%) | 267 (8.2%) | 1867 (7.8%) |

*APOL1* monoallelic, carrying G0G1 or G0G2 genotypes. *APOL1* biallelic, carrying 2 *APOL1* risk variants, either two copies of the *APOL1* G1 risk mutation, two copies of the *APOL1* G2 risk mutation or one copy of *APOL1* G1 and one copy of *APOL1* G2. BP, Blood pressure. eGFR, estimated Glomerular Filtration Rate. CKD, stage 3/stage 4 chronic kidney disease. ESKD, end stage kidney disease. FSGS, Focal Segmental Glomerular Sclerosis.

Consistent with previous reports and other cohorts, 13.6% (n=3,252) of the participants had two APOL1 HR variants [G1/G1: 5.6%, G1/G2: 6.2% and G2/G2: 1.9%]. 44,52% (n=10,622) had one risk variant [G0/G1: 27.8% and G0/G2: 17.8%] and 41.9% (n=9,983) were G0/G0^2, 3^. There were no differences in age, gender or BMI across these genetic strata (**Table 1**).

### Association of APOL1 Risk Variants and CKD (primary outcome)

The proportion of patients affected by CKD was the highest amongst biallelic carriers, at 32.4% (n=1,054), compared with 24.2% (n=2,566) in monoallelic carriers (G1/G0 or G2/G0) and 21.7% (n=2,164) among G0/G0 individuals. End-stage kidney disease was also more frequent in biallelic carriers (12.4% n=403) than the monoallelic carriers (4.5% n=476) or G0/G0 individuals (3.9% n=387).

APOL1 biallelic high-risk carriers had an increased odds of CKD (adjusted odds ratio *a*OR = 1.69; 95% confidence interval [CI], 1.54 to 1.85) compared to the low-risk carriers (< 2 risk alleles) in the minimally adjusted model which adjusted for age, sex, diabetes and the first 3 principal components of ancestry. The *a*OR increased to 1.72 (95% CI, 1.57 to 1.89) in the fully adjusted model, which additionally adjusted for mean arterial pressure, HIV status, body mass index, RAASi use, SGLT2 inhibitor use and the number of antihypertensive drugs (**Table 2**).

**Table 2.** Association of *APOL1* Genotypes with CKD.

| *APOL1* Genotypes | *Odds Ratio (95% CI) |  |  |
| --- | --- | --- | --- |
|  | Unadjusted | Minimally adjusted | Fully adjusted |
| **Primary Outcome: Chronic Kidney Disease** |  |  |  |
| *Sample size 5784/18,073* |  |  |  |
| 2 *APOL1*HR vs. < 2 (recessive) | 1.61 (1.49, 1.74) | 1.69 (1.54, 1.85) | 1.72 (1.57, 1.89) |
| *APOL1* HR additive model | 1.28 (1.23, 1.34) | 1.27 (1.21, 1.33) | 1.28 (1.22, 1.34) |
| *Individual APOL1 genotypes (variable sample size depending on the count for the genotype)* |  |  |  |
| G1/G1 vs. G0/G0 | 2.02 (1.79, 2.28) | 2.08 (1.82, 2.39) | 2.06 (1.79, 2.37) |
| G1/G2 vs. G0/G0 | 1.58 (1.40, 1.78) | 1.56 (1.37, 1.79) | 1.59 (1.38, 1.83) |
| G2/G2 vs. G0/G0 | 1.44(1.17, 1.78) | 1.58 (1.25, 2.00) | 1.64 (1.28, 2.10) |
| G0/G1 and G0/G2 vs. G0/G0 | 1.15 (1.08, 1.23) | 1.08 (1.00, 1.16) | 1.07 (0.99, 1.15) † |
| G0/G2 vs. G0G0 | 1.22 (1.12, 1.33) | 1.12 (1.02, 1.23) | 1.11 (1.01, 1.23) |
| G0/G1 vs. G0/G0 | 1.11 (1.03, 1.20) | 1.05 (0.97, 1.14) † | 1.04 (0.96, 1.13) † |
| **Secondary Outcomes** |  |  |  |
| **Proteinuria** |  |  |  |
| *Sample size 1533/22,324* |  |  |  |
| 2 *APOL1*HR vs. < 2 (recessive) | 2.15 (1.90, 2.43) | 2.02 (1.77, 2.33) | 2.02 (1.77, 2.31) |
| *APOL1* HR additive model | 1.55 (1.45, 1.67) | 1.48 (1.37, 1.60) | 1.48 (1.37, 1.60) |
| *Individual APOL1 genotypes (variable sample size depending on the count for the genotype)* |  |  |  |
| G1/G1 vs. G0/G0 | 2.80 (2.33, 3.37) | 2.43 (2.00, 2.96) | 2.37 (1.94, 2.91) |
| G1/G2 vs. G0/G0 | 2.49 (2.07, 2.99) | 2.28 (1.88, 2.78) | 2.32 (1.90, 2.84) |
| G2/G2 vs. G0/G0 | 1.62 (1.13, 2.32) | 1.58 (1.09, 2.30) | 1.63 (1.11, 2.40) |
| G0/G1 and G0/G2 vs. G0/G0 | 1.32 (1.17, 1.48) | 1.23 (1.09, 1.39) | 1.23 (1.09, 1.39) |
| G0/G2 vs. G0G0 | 1.47 (1.27, 1.71) | 1.37 (1.17, 1.60) | 1.34 (1.15, 1.59) |
| G0/G1 vs. G0/G0 | 1.22 (1.07, 1.40) | 1.15 (1.00, 1.32) † | 1.14 (0.99, 1.32) † |
| **FSGS** |  |  |  |
| *Sample size 80/18,073* |  |  |  |
| 2 *APOL1*HR vs. < 2 (recessive) | 17.67 (10.91, 28.61) | 17.83 (10.76, 29.55) | 17.48 (10.53, 29.02) |
| *APOL1* HR additive model | 7.91 (5.33, 11.73) | 8.07 (5.36, 12.16) | 7.93 (5.25, 11.98) |
| *Individual APOL1 genotypes (variable sample size depending on the count for the genotype)* |  |  |  |
| G1/G1 vs. G0/G0 | 34.77 (15.51, 77.97) | 41.68 (16.67, 104.24) | 35.32 (14.21, 87.83) |
| G1/G2 vs. G0/G0 | 27.88 (12.40, 62.67) | 28.43 (11.92, 67.83) | 30.73 (12.65, 74.67) |
| G0/G1 vs. G0/G0 | 3.19 (1.33, 7.63) | 2.99 (1.25, 7.18) | 2.83 (1.18, 6.79) |

\* odds ratios (95% confidence interval) for the association *APOL1* genotypes with the outcomes.
^ For proteinuria controls are anyone without proteinuria, including those with low GFR.
The primary analysis was adjusted for age, sex, diabetes and first 3 principal components of ancestry. In the secondary analysis we further adjusted for BMI, HIV status, mean arterial pressure, RAASi, number of antihypertensive drugs and sodium glucose (SGLT2i).
For FSGS comparison analyses were completed using firth logistic regression. Association for FSGS and G0/G2 or G2G2 were not conducted given 1 case with G0/G2 and 2 cases with G2/G2
† Not statistically significant

A modest association between monoallelic APOL1 risk variants carriers and CKD was also observed. Participants with a single risk allele (genotype G0/G1 or G0/G2) had higher odds of having CKD (*a*OR = 1.08; 95% CI, 1.00 to 1.16; p=0.04) in the minimally adjusted model (**Table 2**). This was driven by the G0/G2 vs G0/G0 (*a*OR = 1.11; 95% CI, 1.01 to 1.23; p=0.03). No significant association was observed in the fully adjusted model or for G0/G1 vs G0/G0.

### Association of APOL1 risk variants with proteinuria, and FSGS (secondary outcome)

#### Association with main proteinuria outcome

the main proteinuria outcome was peak protein creatinine ratio >700 mg/g or albumin to creatinine ratio > 420 mg/g, which was reached by 1,533 participants. This cut-off was chosen based on the Inaxaplin phase 2 study to further inform the effect of allelic dosing in proteinuric CKD^13^. The proportion of patients with PCR >700 mg/g was higher among the biallelic 370 (11.4%) and monoallelic 674 (6.3%), and the lowest for G0/G0 489 (4.9%).

APOL1 biallelic high-risk carriers had an increased odds of proteinuria (*a*OR = 2.02; 95% CI, 1.77 to 2.33, p<0.001) compared to the low risk carriers (< 2 risk alleles), adjusting for age, sex, diabetes status and the first top 3 principal components of ancestry. In the fully adjusted model, the *aOR* for this association remained same (*a*OR = 2.02; 95% CI, 1.77 to 2.31) (**Table 2**). Sequential models also showed an association between APOL1 monoallelic risk variants carriers (G0/G1 plus G0/G2 versus G0/G0) and proteinuria (*aOR* = 1.23; 95% CI, 1.09 to 1.39) in the minimally adjusted model. The fully adjusted model had nearly identical estimates (*aOR* = 1.23; 95% CI, 1.09 to 1.39). This supports the strongest association with the biallelic, and moderate association with monoallelic and proteinuria compared to G0/G0.

#### Associations with FSGS

Eighty patients had FSGS, 10 of them had collaping FSGS and five had biopsies that showed complete podocyte foot effacement, and given the known risk of sampling error were treated as FSGS. Collapsing FSGS was seen only in Biallelic carriers. Overall 71.3% of the FSGS cases were in biallelic carriers, 20.0% in monoallelic carriers and 8.8% in G0/G0. The predominant risk allele was G1, there was only one patient that was G0/G2 and two patient that were G2/G2 (**Figure 2**).

**Figure 2.**
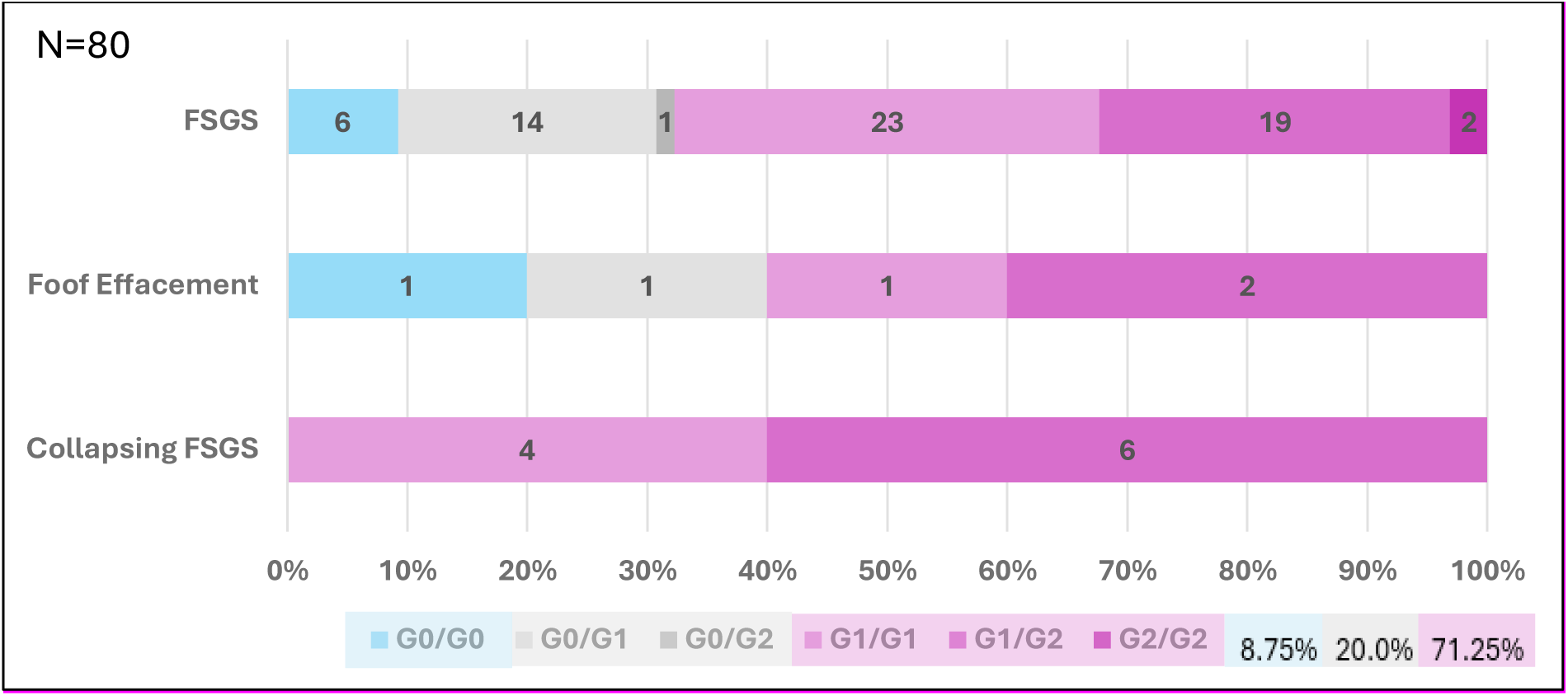
APOL1 genotype count by FSGS category

There was an association for the biallelic and FSGS (2 APOL1HR vs. < 2 (recessive) (*aOR* =17.83, 95%, CI 10.76 to 29.55). We also observed an APOL1 dosing effect in the additive model of inheritance (*aOR=* 8.07,95% CI 5.36 to 12.16). There was no independent analysis that included G0/G2 or G2/G2 due to the scarcity of these genotypes. The only monoallelic comparisons were G0/G1 versus G0/G0 (*aOR* =2.99, (95%CI 1.25 to 7.18). The strongest association was for G1/G1 versus G0/G0 (*aOR* =41.68, 95% CI 16.67 to 104.24). For G1/G2 vs G0/G0 *aOR* was 28.43 (95% CI 11.92, 67.83).

### Sensitivity Analyses

#### Protein to creatinine ratio as a continuous measurement

The mean highest PCR value among those with measurements (n=4050) was 2.24 g/g (SD 3.99) amongst biallelic carriers and 1.59 g/g (SD 3.15) amongst monoallelic carriers compared to 1.27 g/g (SD 2.63) for patients with G0/G0. There was an association between both the biallelic and the monoallelic APOL1 carriers and the highest protein creatinine ratio value in the 4055 patients that had a measurement available. For the minimally adjusted model with log transformed highest PCR, the biallelic carriers showed 1.59 times (95% CI: 1.40 to 1.79; p < 0.0001) highest PCR value versus patients with G0/G0, and monoallelic carriers showed 1.13 times (95% CI: 1.01 to 1.25; p = 0.03) highest PCR value versus patients with G0/G0 (**Figure 3**).

**Figure 3.**
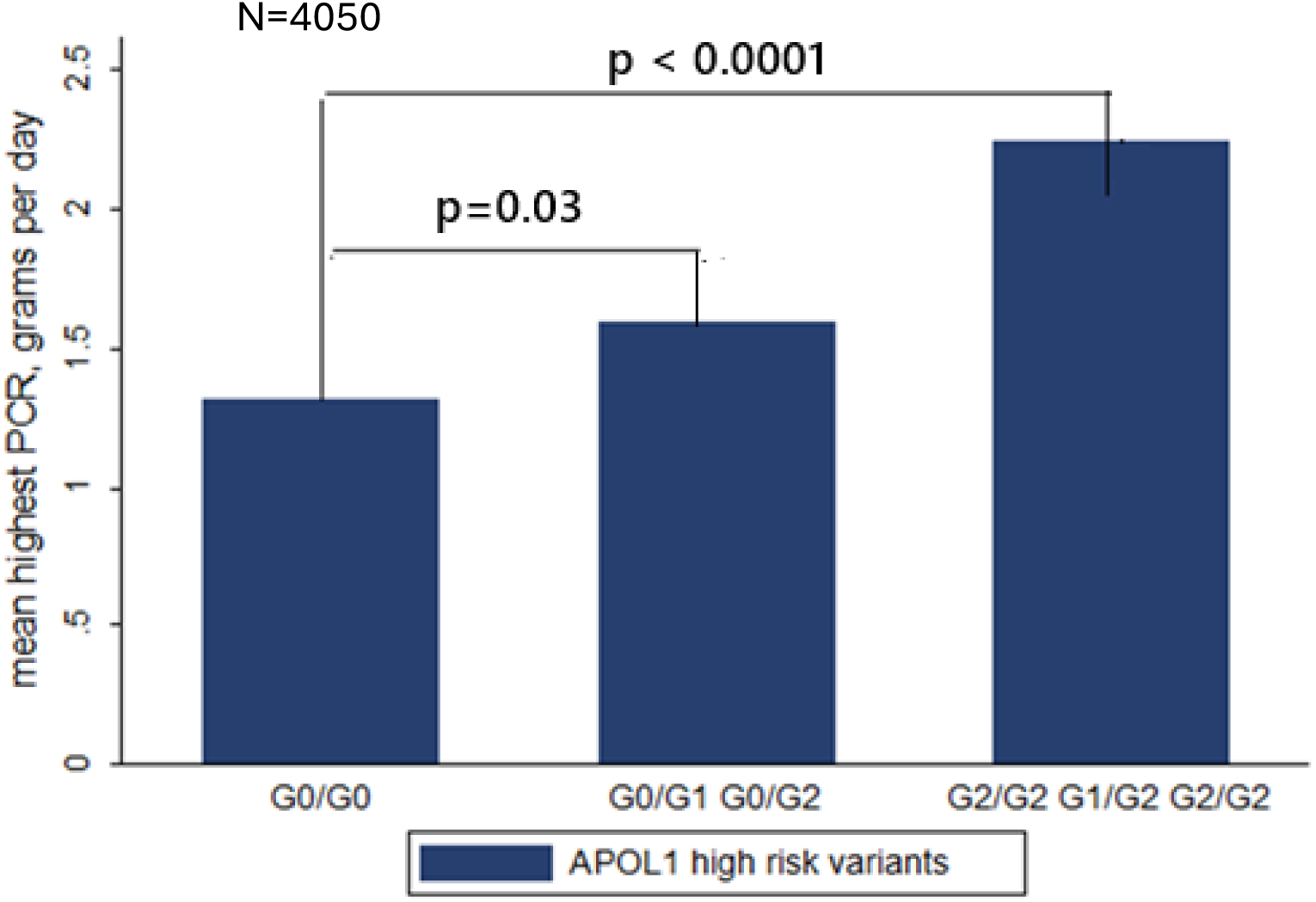
Association of APOL1 Monoallelic and Biallelic with the highest PCR value recorded in all with protein creatinine ratio measurements (n=4,050). **\***Linear regression of log transformed PCR adjusted for age, sex, dm and 3 PCs of ancestry

#### Additive model of inheritance

This model was consistent with an APOL1 risk variants allelic dosing or graded association with CKD, minimally adjusted model (aOR=1.27,95% CI, 1.21 to 1.33) and fully adjusted model 1.28 (95% CI 1.22 to 1.34). In the additive model there were significant associations with proteinuria with similar results in both the minimally and fully adjusted models (aOR =1.48, 95%CI, 1.37 to 1.60), as well as FSGS (aOR=8.07, 95%CI 5.36 to 12.16) in the minimally adjusted and aOR=7.93 (5.25 to 11.98) in the fully adjusted model.

## Discussion

Our study reports that in African ancestry participants from the Vanderbilt University Medical Center Biobank, monoallelic APOL1 genotypes were associated with 8% higher odds of CKD, 23% higher odds of proteinuria and 3-fold higher odds of FSGS for G0/G1. Biallelic APOL1 genotypes had a stronger association with all kidney phenotypes as expected. This study add to the evidence provided by the study in the West African population by confirming the association in an US based cohort of monoallelic APOL1 genotypes and AMKD and by providing the graded association with proteinuria.

The association of monoallelic genotypes and CKD was modest in our study (8%) compared to the one observed in H3Africa (18% increase odds for CKD). This may be in part due to disease heterogeneity in our population. However, the additive model of inheritance showed that APOL1 variants have an allelic dosing effect and a graded association with all kidney outcomes: CKD, proteinuria and FSGS.

In our study we evaluated the association with proteinuria using two different approaches. A cutoff of 700 mg/g and what was considered an equivalent in urine albumin to creatinine ratio of 420 mg/g. This was chosen based in the cut off describe in the PARASOL and replicated in RaDaR study^14^ . Describing the association with this cutoff was important as real world evidence anlaysis from PARASOL and RaDaR have supported that dropping proteinuria below 700 mg/g had signifcant lower hazard of kidney failure and other meaningful clinical outcomes and have provided the bases as a clinical trial endpoint for an accelerated approval for drugs for rare disease like FSGS^12^. This is important for AMKD as clinical trials have many barriers in completing their enrollment particularly because the low number of patients that meet criteria. In our study both monoallelic and biallelic APOL1 genotypes demonstrated associations with proteinuria exceeding the 700 mg/g cutoff value. This warrants further investigation in larger and longitudinal cohorts, particularly as individuals with these genotypes may be candidates for APOL1 targeted therapies. The second approach used in the study was using the highest PCR for all the patients with a measurement available. This approach allowed us to study associations acros the entire range of proteinuria values, including lower values that may not be captured by fixed cutoffs. and support that studies at different cutoff also need to be pursued as there is an allelic dosing effect and graded association with APOL1 risk alleles. The associations with lower levels of proteinuria are likely to be seen in longer peirods of follow up, which could be troublesome for clinical trials but that can be capture by real world evidence analysis.

Our study only had 80 patients with biopsy proven-FSGS, from whom 10 had collapsing FSGS. Collaping FSGS was seen only in biallelic carrier in this study (G1/G1 or G1/G2). It is important to highlight that collapsing features have been described in monoallelic carrier with untreated HIV^7^ and severe COVID-19^15^, supporting the role of enviromental factors and their interaction as an important driver of the APOL1 mediated kidney phenotype. For all FSGS, 70% were observed in biallelic carriers, 20% in monoallelics and only 7 cases were G0/G0. FSGS was very uncommon in a G2 genotype (3 cases total). There was an allelic dosing and association with a monoallelic for G1/G0 with an odds ratio of 7 to 8.

Our study must be interpreted considering limitations. Our study represents a real-world evidence analysis where data is collected as part of routine clinical care. However, our results are consistent with those from the West Africa cohort^8^. Another limitation of our study was the small number of biopsy-proven FSGS cases at this time. Strenghts of our cohort include its generalizability, as the prevalence for biallelic (homozygous or compound heterozygous) and monoalleic (heterozygous) APOL1 genotypes is similar to other US-based cohorts^2, 3, 16^. The comprehensive AGD/BioVU dataset also provided a wealth of phenotypic information and allowed for a detailed assessment between APOL1 genotypes and clinical outcomes based on patient electronic health records.

In conclusion, both monoallelic carriers and biallelic carriers had increased odds of CKD, proteinuria, both at a cutoff of 700 mg/g and as a continuous variable, and FSGS in this US-based cohort. Participants with one risk allele had an 8% increase in the odds of CKD, 23% increase in the odds of proteinuria (>700 mg/g) and 3-fold increase odds for FSGS (G1/G0 only). Furthermore, the additive model of inheritance showed an allelic dosing effect for these outcomes. Our study confirms the associations described in the West Africa cohort and provides additional associations between monoallelic variants and proteinuria. Larger and longitudinal studies in human populations are needed to further understand genes, haplotype background and environmental interactions that further modify the risk of disease progression in both those with 1 risk alleles and those with 2 risk alleles. This will inform further decisions on personalized care, particularly in the treatment approach for monoallelic carriers with clinical manifestations of kidney disease, as well as to inform the proteinuria cutoff at which targeted interventions should be considered.

## Data Availability

The data belongs to BioVu, the Vanderbilt Biobank, and the data is not publicly available.

## Acknowledgment

We are grateful to the AGD/BioVU Pilot project and QC team for the generation of the whole genome data for the AGD linked to Vanderbilt electronic medical center and to the big data team for procesing of the synthetic derivative. A.M.H. had full access to all the data in the study and assumed responsibility for data integrity. The project described was supported by CTSA award No. UL1 TR002243 from the National Center for Advancing Translational Sciences. Its contents are solely the responsibility of the authors and do not necessarily represent official views of the National Center for Advancing Translational Sciences or the National Institutes of Health.

## Supplemental information

**Supplemental table 1.** Comorbidities definition.

| **Dialysis** |  |
| --- | --- |
| ICD-9<br>Diagnosis | V45.1, V45.11, V56.0, V56.1, V56.2, V56.8, 585.6, V45.12, V56.31, V56.32, 792.5, 39.95, 39.952, 54.982, 39.953, 54.98, 54.983, 99.78 |
| ICD-10<br>diagnosis | N18.6 R88.0 Z49.31, Z49.32, Z91.15, 3E1M39Z Z49.01, Z49.02, Z99.2 |
| ICD9 PC | 39.951, 54.981 |
| CPT | 90941, 90942, 90943, 90944, G0491, G0492, M0916, M0920, M0931, M0932, 90996, A4690, A4750, A4755, 90935, 90937, 90947, 90999, 90988, 90997, 4054F, 4055F, E1590, G8715, G8727, M0936, 36516, 36901, 36902, 36903, 36904, 36905, 36906, 36516, 36901, 36902, 36903, 36904, 36905, 36906, 36907, 36908, 36909, 4052F, 4053F, 90939, 90940, 90963, 90964, 90965, 90966, 90967, 90968, 90969, 90970, 90976, 90977, 90978, 90979, 90982, 90983, 90984, 90985, 90991, 90994, 99512, 99559, G0257, G0320, G0321, G0322, G0323, G0327, G8075, G8076, G8714, G8956, G9264, G9265, G9266, M0923, M0928, M0937, M0940, M0944, M0945, M0948, M0952, M0956, S9335, S9339, G9240, G9241, 0505F, 0507F, 9092, 90921, 90924, 90925, 90951, 90952, 90953, 90954, 90955, 90956, 90957, 90958, 90959, 90960, 90961, 90962, 90989, 90990, 90992, 90993, 90995, 90998, 4054F, 4055F |
| ICD-10 | E11.00, E11.01, E11.10, E11.11, E11.21, E11.22, E11.29, E11.311, E11.319, E11.321, E11.3211, E11.3212, E11.3213, E11.3219, E11.329, E11.3291, E11.3292, E11.3293, E11.3299, E11.331, E11.3311, E11.3312, E11.3313, E11.3319, E11.339, E11.3391, E11.3392, E11.3393, E11.3399, E11.341, E11.3411, E11.3412, E11.3413, E11.3419, E11.349, E11.3491, E11.3492, E11.3493, E11.3499, E11.351, E11.3511, E11.3512, E11.3513, E11.3519, E11.3521, E11.3522, E11.3523, E11.3529, E11.3531, E11.3532, E11.3533, E11.3539, E11.3541, E11.3542, E11.3543, E11.3549, E11.3551, E11.3552, E11.3553, E11.3559, E11.359, E11.3591, E11.3592, E11.3593, E11.3599, E11.36, E11.37X1, E11.37X2, E11.37X3, E11.37X9, E11.39, E11.40, E11.41, E11.42, E11.43, E11.44, E11.49, E11.51, E11.52, E11.59, E11.610, E11.618, E11.620, E11.621, E11.622, E11.628, E11.630, E11.638, E11.641, E11.649, E11.65, E11.69, E11.8, E11.9, O24.111, O24.112, O24.113, O24.119, O24.12, O24.13 |
| ICD9 | 250, 250.1, 250.2, 250.3, 250.4, 250.5, 250.6, 250.7, 250.8, 250.9, 357.2, 362.01, 362.02, 362.04, 362.05, 362.06, 362.07, 366.41, 648, 648.01, 648.03, 648.04 |
| **Hypertension** |  |
| ICD10 | I10., O10.011, O10.012, O10.013, O10.019, O10.02, O10.03, O10.911, O10.912, O10.913, O10.919, O10.92, O10.93, O16.1, O16.2, O16.3, O16.4, O16.5, O16.9 |
| ICD-9 | 401,401.1,401.9,402,402.01,402.1,402.11,402.9,402.91,403,403.01,403.1,403.11, 403.9,403.91,404, 404.01, 404.02, 404.03, 404.1, 404.11, 404.12, 404.13, 404.9, 404.91, 404.92, 404.93, 405.01, 405.09, 405.11, 405.19, 405.91, 405.99, 437.2, 642, 642.01, 642.02, 642.03, 642.04, 642.1, 642.11, 642.12, 642.13, 642.14, 642.2, 642.21, 642.22, 642.23, 642.24, 642.3, 642.31, 642.32, 642.33, 642.34, 642.4, 642.41, 642.42, 642.43, 642.44, 642.5, 642.51, 642.52, 642.53, 642.54, 642.6, 642.61, 642.62, 642.63, 642.64, 642.7, 642.71, 642.72, 642.73, 642.74, 642.9, 642.91, 642.92, 642.93 |
| Nephrotic Syndrome |  |
| ICD10 | N04.0, N04.1, N04.2, N04.3, N04.4, N04.5, N04.6, N04.7, N04.8, N04.9 |
| ICD9 | 581.9 |
| FSGS |  |
| ICD10 | Narrow N04.1<br>Broad: N00.1, N01.1, N02.1 N03.1 N04.1 N05.1, N06.1, N07.1, N08 |
| **Alport's** |  |
| ICD10 | Q87.81 |
| ICD9 | 759.89 |
| **ADPKD** |  |
| ICD10 | Q61.2, Q61.3 Z82.71 |
| ICD9 | 753.12, 753.13 |
| **DKD** |  |
| ICD-9 | <u>250.40, 250.41, 250.42, 250.43,</u> |
| ICD-10 | E11.22 |
| **CKD** |  |
| ICD-9 | <u>250.40, 250.41, 250.42, 250.43, 403.9, 403.90, 403.91, 582, 582.1, 582.2, 582.4, 582.81, 582.89, 582.9, 585., 585.1, 585.2, 585.3, 585.4, 585.5, 585.6, 585.9, 75</u> |
| ICD-10 | <u>E11.22, I12.9, N03.0, N03.1, N03.2, N03.3, N03.4, N03.5, N03.6, N03.7, N03.8, N03.9, N03.A, N18.1, N18.2, N18.3, N18.30, N18.31, N18.32, N18.4, N18.5, N18.</u> |
| Kidney transplant |  |
| ICD-9 | V42.0, 55.69 |
| ICD-10 | Z94.0, Z48.22, T86.11, 0TY00Z0 0TY00Z1 0TY00Z2 0TY10Z0 0TY10Z1 0TY10Z2 |
| CPT | 50360 50365 |
| **Diabetes** |  |
| ICD-9 | <u>250.00, 250.01, 250.02, 250.03, 250.10, 250.11, 250.13, 250.20, 250.21, 250.22, 250.23, 250.31, 250.32, 250.40, 250.41, 250.50, 250.52, 250.60, 250.61, 250.63, 250.70, 250.71, 250.72, 250.73</u> |
| ICD-10 | E08.00, E08.01, E08.22, E08.29, E08.311, E08.319, E08.321, E08.3211, E08.3212, E08.3219, E08.329, E08.3293, E08.3312, E08.339, E08.3391, E08.3392, E08.3393, E08.349, E08.3492, E08.3499, E08.3511, E08.3513, E08.3519, E08.3521, E08.3522, E08.3523, E08.3532, E08.3533, E08.3542, E08.3549, E09.00, E09.01, E09.10, E09.11, E09.319, E09.3211, E09.3213, E09.3291, E09.3292, E09.331, E09.3311, E09.3313, E09.3391, E09.3393, E09.3399, E09.3411, E09.3419, E09.3493, E09.3499, E09.3512, E09.3521, E09.3522, E09.3529, E09.3531, E09.3532, E09.3549, E09.3593, E09.37X1, E09.37X2, E09.37X9, E10.10, E10.21, E10.22, E10.29, E10.311, E10.3213, E10.3219, E10.3292, E10.3293, E10.3299, E10.3311, E10.3391, E10.3393, E10.3411, E10.3412, E10.3419, E10.3493, E10.351, E10.3511, E10.3512, E10.3521, E10.3523, E10.3533, E10.3542, E10.3543, E10.3551, E10.3553, E10.3559, E10.359, E10.3592, E11.00, E11.10, E11.11, E11.22, E11.311, E11.319, E11.3211, E11.3212, E11.3213, E11.3219, E11.3291, E11.3299, E11.3391, E11.3393, E11.3412, E11.3419, E11.349, E11.3491, E11.3499, E11.351, E11.3513, E11.3519, E11.3521, E11.3529, E11.3531, E11.3539, E11.3542, E11.3551, E11.3552, E11.3553, E13.10, E13.11, E13.21, E13.22, E13.29, E13.319, E13.321, E13.3212, E13.3219, E13.329, E13.3291, E13.3293, E13.3299, E13.3311, E13.3313, E13.3319, E13.3393, E13.3399, E13.341, E13.349, E13.3492, E13.3493, E13.3499, E13.3511, E13.3512, E13.3519, E13.3521, E13.3523, E13.3529, E13.3539 |
| Medications | ACARBOSE, ALBIGLUTIDE, ALOGLIPTIN, ALOGLIPTIN/<br>METFORMIN, ALOGLIPTIN/PIOGLITAZONE, CANAGLIFLOZIN, CANAGLIFLOZIN/<br>METFORMIN, CHLORPROPAMIDE, DAPAGLIFLOZIN, DAPAGLIFLOZIN/<br>METFORMIN, DULAGLUTIDE, EMPAGLIFLOZIN, EMPAGLIFLOZIN/<br>LINAGLIPTIN, EMPAGLIFLOZIN/<br>METFORMIN, EXENATIDE, GLIMEPIRIDE, GLIMEPIRIDE/<br>PIOGLITAZONE, GLIMEPIRIDE/ROSIGLITAZONE, GLIPIZIDE, GLIPIZIDE/<br>METFORMIN, GLYBURIDE, GLYBURIDE/METFORMIN, INSULIN -<br>BASAL, INSULIN - OTHER, INSULIN - PREMIXED, INSULIN -<br>SHORT, LINAGLIPTIN, LINAGLIPTIN/<br>METFORMIN, LIRAGLUTIDE, LIXISENATIDE, METFORMIN, METFORMIN/<br>PIOGLITAZONE, METFORMIN/REPAGLINIDE, METFORMIN/<br>ROSIGLITAZONE, METFORMIN/SAXAGLIPTIN, METFORMIN/<br>SITAGLIPTIN, NATEGLINIDE, PIOGLITAZONE, PIOGLITAZONE/<br>METFORMIN, PRAMLINTIDE, REPAGLINIDE, ROSIGLITAZONE, ROSIGLITAZONE/<br>GLIMEPIRIDE, ROSIGLITAZONE/METFORMIN, SAXAGLIPTIN, SIMVASTATIN/<br>SITAGLIPTIN, SITAGLIPTIN, TOLAZAMIDE, TOLBUTAMIDE, TROGLITAZONE |
| **Sickle cell Disease: by code or by being homozygous for rs334-a** |  |
|  | Contains ICD10 code D57.00-Hb-SS disease with crisis, unspecified<br>Contains ICD10 code D57.01-Hb-SS disease with acute chest syndrome<br>Contains ICD10 code D57.02-Hb-SS disease with splenic sequestration<br>Contains ICD10 code D57.1-Sickle-cell disease without crisis<br>Contains ICD10 code D57.3-Sickle-cell trait<br>Contains ICD9 code 282.41-Sickle-cell thalassemia without crisis<br>Contains ICD9 code 282.6-Sickle-cell disease |
|  | Contains ICD9 code 282.5-Sickle-cell trait<br>Contains ICD9 code 282.42-Sickle-cell thalassemia with crisis<br>Contains ICD9 code 282.60-Sickle-cell disease, unspecified<br>Contains ICD9 code 282.69-Other sickle-cell disease with crisis<br>Contains ICD9 code 282.68-Other sickle-cell disease without crisis<br>Contains ICD9 code 282.61-Hb-SS disease without crisis<br>Contains ICD9 code 282.62-Hb-SS disease with crisis |
| **Lung transplant** |  |
| ICD-9 | 996.84, V42.6 |
| ICD-10 | T86.81, T86.810, T86.811, T86.812, T86.818, T86.819, Z48.24, Z48.280, Z94.2, Z94.3 |
| **Heart transplant** |  |
|  | 414.06, 414.07, 429.4, 996.83, V15.1, V42.1, V43.2, V43.21, V43.22 |
|  | I25.750, I25.751, I25.758, I25.759, I25.760, I25.761, I25.768, I25.769, I25.811, I25.812, I97.0, I97.110, I97.111, I97.120, I97.121, I97.130, I97.131, I97.190, I97.191, T86.2, T86.20, T86.21, T86.22, T86.23, T86.290, T86.298, T86.30, T86.31, T86.32, T86.33, T86.39, Z48.21, Z48.280, Z94.1, Z94.3, Z95.811, Z95.812 |
| **Complicated cirrhosis** |  |
|  | 571.2, 571.5, 571.6 plus 789.5, 789.59 R18, R18.8 |

